# Direct and indirect impact of the COVID-19 pandemic on the survival of kidney transplant recipients: a national observational study in France

**DOI:** 10.1101/2023.04.05.23288113

**Authors:** Elhadji Leye, Tristan Delory, Khalil El Karoui, Maude Espagnacq, Myriam Khlat, Sophie Le Coeur, Nathanaël Lapidus, Gilles Hejblum, the COVID-HOSP working group

## Abstract

**Background:** During the pandemic period, healthcare systems were substantially reorganized for managing COVID-19 cases. The corresponding changes on the standard care of persons with chronic diseases and the potential consequences on their outcomes remain insufficiently documented. This observational study investigates the direct and indirect impact of the pandemic period on the survival of kidney transplant recipients (KTR), in particular in those not hospitalized for COVID-19.

**Methods:** We conducted a cohort study using the French national health data system which contains all healthcare consumptions in France. Incident persons with end stage kidney disease between January 1, 2015 and December 31, 2020 who received a kidney transplant were included and followed-up from their transplantation date to December 31, 2021. The survival of KTR during the pre-pandemic and pandemic periods was investigated using Cox models with time-dependent covariates, including vaccination and hospitalization events.

**Findings:** There were 10,637 KTR included in the study, with 324 and 430 deaths observed during the pre-pandemic (15,115 person-years of follow-up) and pandemic periods (14,657 person-years of follow-up), including 127 deaths observed among the 659 persons with a COVID-19-related hospitalization. In multivariable analyses, the risk of death during the pandemic period was similar to that observed during the pre-pandemic period (hazard ratio (HR) [95% confidence interval]: 0 ·92 [0·77–1·11]), while COVID-19-related hospitalization was associated with an increased risk of death (HR: 10 ·62 [8·46–13·33]). In addition, pre-emptive kidney transplantation was associated with a lower risk of death (HR: 0·71 [0·56–0·89]), as well as a third vaccine dose (HR: 0·42 [0·30– 0·57]), while age, diabetes and cardiovascular diseases were associated with higher risks of death.

**Interpretation:** Considering persons living with a kidney transplant with no severe COVID-19-related hospitalization, the pandemic period was not associated with a higher risk of death.

**Funding:** Initiative Économie de la Santé de Sorbonne Université (Idex Sorbonne Université, programmes Investissements d’Avenir); Ministère de la Solidarité et de la Santé (PREPS 20-0163).

## Introduction

Kidney transplant recipients (KTR) are at increased susceptibility to many viral infections because of their immunosuppressive treatments, and this led to justifiable anxiety about the effect of COVID-19 on them.^1,2^ As mentioned by Vart et al, ^3^ KTR were quickly identified as a vulnerable population, prompting tremendous efforts from the nephrology community to gather data informing clinical practice in record time. Reviews examined princeps studies which collected and reported such data on KTR, ^4–7^ and indicate that documenting the direct impact of COVID-19 in KTR raised many concerns. The issues addressed evolved with time and may be roughly categorized according to whether these studies were conducted before or after the availability of vaccines against COVID-19. Before vaccine availability, a major concern was the mortality of KTR with COVID-19, and a review based on 74 studies totalizing 5,559 KTR estimated this risk at 23% [95% confidence interval: 21%–27%].^5^ A study based on the 87,809 hospital admissions with COVID-19 that occurred in France until June 15, 2020, reported that compared to the general population, KTR had a 4 ·6-fold higher risk of being hospitalized with COVID-19, and a 7·1-fold higher risk of in-hospital mortality.^8^ After vaccination roll-out, a major concern was assessing the protection provided by the vaccines, ^7^ including a personalization of the vaccination schedule in KTR in order to take into account their decreased immune response. ^9,10^ A French national study conducted in the earliest period of the vaccination period, when two doses constituted a complete vaccination schedule, reported that compared to the persons vaccinated in the general population, vaccinated KTR had a 5 ·9-fold higher risk of hospitalization for COVID-19 and a 6·3-fold higher risk of in-hospital mortality. ^11^

In contrast with the great interest brought to documenting the direct impact of COVID-19 on KTR, the indirect impact of the pandemic on the health of KTR is a more complex topic to investigate. More precisely, although most KTR did not experience a severe episode of COVID-19, to our knowledge no study has yet addressed the following important issue: were the pandemic-related changes in healthcare organization simultaneously associated with a higher mortality in KTR who did not experience a severe episode of COVID-19? Indeed, in many countries, healthcare systems were reorganized during the COVID-19 pandemic in order to manage symptomatic cases of COVID-19 and such a reorganization may have indirectly impacted the health outcomes of those who did not experience a severe COVID-19 episode but who suffered from other diseases, particularly persons with chronic diseases including KTR. For example, in France, intensive care capacity was prioritized and extended during the pandemic, with a maximum of 10,000 beds available while the national capacity of such beds was 5,080 before the pandemic.^12^ Conversely, the activity of kidney transplantation was totally suspended between March 10 and May 15, 2020.^13^

In order to investigate the indirect impact of the pandemic on KTR, we undertook a national observational study in France comparing the survival (all-cause mortality) in this vulnerable population before and during the pandemic. Our main goal was to investigate if there was an excess mortality in KTR not hospitalized with COVID-19 during the pandemic. The longitudinal analyses performed took into account the potential role of relevant covariates such as comorbidities, age, time between the initiation of dialysis and the transplantation date, and time since transplantation date. Assessing the indirect impact of the pandemic on the survival of KTR also required estimating the direct effect of COVID-19. Therefore, this study also details the dynamics of COVID-19 vaccination in KTR in France, and investigates how vaccination may have impacted COVID-19-related hospitalizations and survival in KTR.

## Methods

This observational study was conducted according to STROBE guidelines. ^14^

This study was based on data from the Système National des Données de Santé (SNDS) Database, a French National Health Data System which covers nearly 99% of the French population. ^15^ Any health care consumption subjected to a reimbursement by the national medical insurance system is recorded and linked to the individual who received the corresponding healthcare through a unique and anonymous identifier. Therefore, data include prescription-based medication deliveries with corresponding Anatomical Therapeutic Chemical (ATC) codes and dates of delivery, medical consultations, hospitalizations with corresponding International Classification of Diseases tenth Revision ICD10 codes for primary and related diagnoses, and dates of admission and discharge. The SNDS also contains demographic data, e.g., sex and dates of birth and death, and the date of each dose of COVID-19 vaccine received by a given individual. With the growing interest of the scientific medical community in real-world data from large administrative healthcare databases, the SNDS is increasingly used for pharmacoepidemiologic studies, ^16,17^ including studies on COVID-19.^8,11,18,19^

The study took advantage of the G9 mapping, a useful tool provided within the SNDS which allows an automatic selection of individuals labelled with a given chronic disease in a given year. ^20^ Several pathologies have been considered in the G9 mapping including persons with end stage terminal kidney disease (ESKD) and we used it for selecting the persons included in the study. This selection is therefore reproducible and based on an algorithm developed by experts of the SNDS and which was extended to all persons in the database from 2015 and onwards.

### Patients included in the study

First, prevalent persons with ESKD during any year between 2015 and 2020 were selected in the SNDS using G9 mapping. These included any individual who had spent more than 45 days on haemodialysis or had a peritoneal dialysis or had benefited from a kidney transplant or was followed-up after transplantation. Then, among this population of prevalent persons with ESKD between 2015 and 2020, we only selected those who developed ESKD during this period (incident cases), benefited from a kidney transplantation before December 31, 2021, and were aged between 18 and 85 years at the date of transplantation. This restriction to incident persons with a transplantation is a critical characteristic of the study, and was adopted in order to guarantee that the history of the disease was duly documented for each individual included in the study, especially the delay between ESKD onset and transplantation date.

### Pre-pandemic and pandemic periods of the study

We set the beginning of the pandemic period on March 1, 2020: the number of nationally reported cases rose from 100 to 1000 between February 29 and March 8, with a first lockdown starting on March 17. Pre-pandemic period was therefore defined between January 1, 2015 and February 29, 2020. The pandemic period was censored on December 31, 2021 considered as the end date of the study. In France, the roll-out of COVID-19 vaccination began on December 27, 2020.

### Statistical analyses

The date of transplantation was considered as the initial time (t_0_) of the corresponding individual follow-up. The investigations on KTR survival during the pre-pandemic and pandemic periods also considered the following additional covariates that we deemed important to simultaneously study: COVID-19-related hospitalization event(s) (COVID-19 as the principal or related code of an hospitalization was considered as a proxy for a severe COVID-19 episode), vaccination against COVID-19, diabetes and cardiovascular comorbidities (including stroke sequelae, chronic heart failure, coronary diseases, peripheral arterial obliterative disease), time spent on dialysis before transplantation date, mode of entry into transplantation (pre-emptive, i.e., without prior dialysis, or not), age and sex. We also explored the effect of the vaccine for preventing COVID-19-related hospitalization. The value of some covariates might have varied along the longitudinal follow-up of the individuals, and accordingly, the following variables were appropriately handled as time-dependent variables in the analyses: period (pandemic versus pre-pandemic), diabetes and cardiovascular comorbidities (baseline presence or incidence versus absence), age, history of COVID-19-related hospitalization, and COVID-19 vaccination. The age variable was transformed into an ordinal variable handled according to the following breakdown: 18–44 (reference), 45–54, 55–64, 65–74 and 75–84 years. COVID-19-related hospitalization was also considered as an ordinal variable with 3 modalities: 0, 1, and ≥2 hospitalizations. Analyses assumed that no information was missing in the database.

Analyses of KTR survival during the pre-pandemic and pandemic periods were conducted using a Cox modelling approach with time-dependent covariates. Therefore, depending on the timing of event and censoring features, some KTR had their period status (either pre-pandemic or pandemic) changed along their follow-up, while other did not (Figure 1). In addition, the study design required censoring the follow-up duration of individuals at five years because no counterfactual observation could be observed for longer durations of follow-up. Several statistical models were fitted: a univariable model (with no covariate), a multivariable model adjusted only for COVID-19-related hospitalizations, and finally a model adjusted for all covariates except vaccination. Additional analyses detailed the schedule of vaccination doses received by the individuals included in this study, and also investigated the relationship between this vaccination and two events: hospitalization for COVID-19 and death.

**Figure 1.**
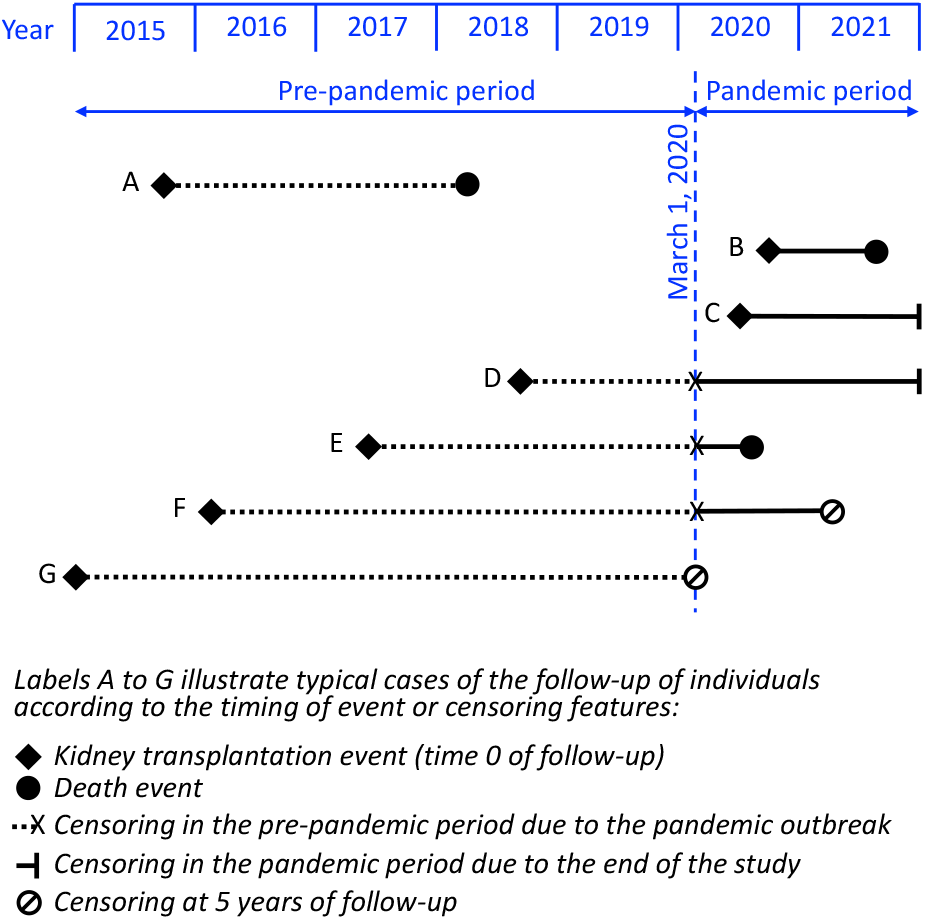
Graphic examples of some individual follow-up in the pre-pandemic and pandemic periods according to the timing of event or censoring features. A, B, and C: whole follow-up of the person in either the pre-pandemic (person A) or the pandemic (person B or C) period; D,E,F: whenever a patient had a transplantation date during the pre-pandemic period and survived until the beginning of the pandemic period, this person left the pre-pandemic follow-up group and entered the pandemic group when the pandemic period began; F and G: follow-up of any person included in the study has been censored at five years since there would not be any counterfactual observation of a person with a follow-up duration greater than five years and two months.

Analyses were performed using R version 4·1·2. A P value < 0·05 was considered statistically significant.

## Results

Figure 2 details the study profile: considering the whole population of adult persons living with ESKD in France each year, from 2015 to 2020 (prevalent cases), the selection of incident ESKD cases occurring each year resulted in a total of 62,827 persons, with eventually 10,637 of them (nearly 17%) who benefited from a first kidney transplant at an age < 85 years old between 2015 to 2021. The inclusion of these 10,637 persons in the study resulted in a total follow-up of 29,772 person-years analysed in this study.

**Figure 2.**
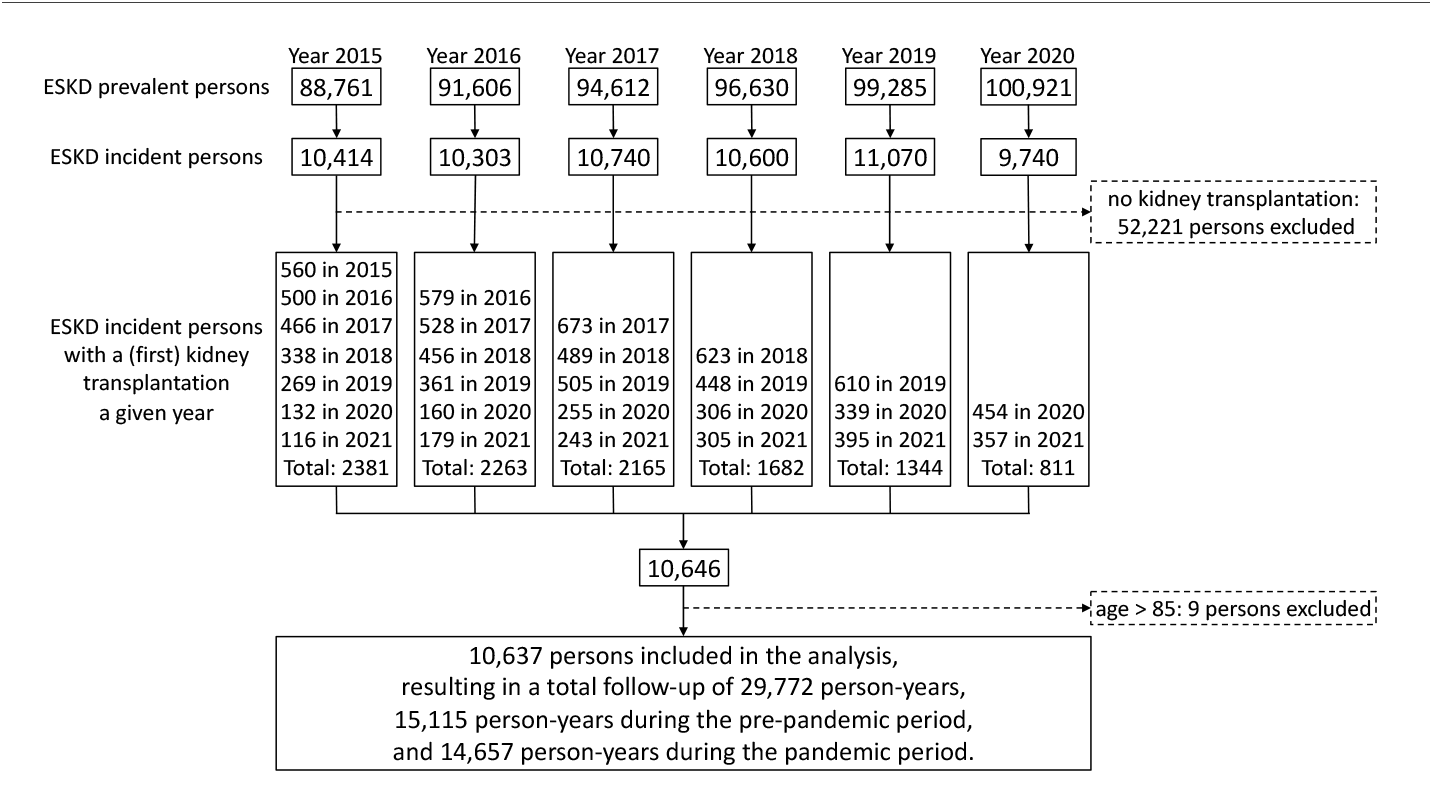
Study flow chart.

Table 1 presents the characteristics of the KTR included in the study at the time of transplantation (baseline). The median age at kidney transplantation was 54 years old (interquartile range (IQR): 43–66), 54 years old (IQR: 42– 65) and 56 years old (IQR: 45–67) for the KTR whose transplantation date was within the pre-pandemic period and the pandemic period, respectively; 64% were males and 44% presented with a comorbidity (diabetes, a chronic cardiovascular disease, or both). The median duration of pre-transplant dialysis was 1 ·21 year (IQR: 0·18–2·33), 0·89 year (IQR: 0·00–1·87) and 2·24 years (IQR: 1·15–3·62) for the KTR whose transplantation date was within the pre-pandemic and pandemic period, respectively.

**Table 1.**
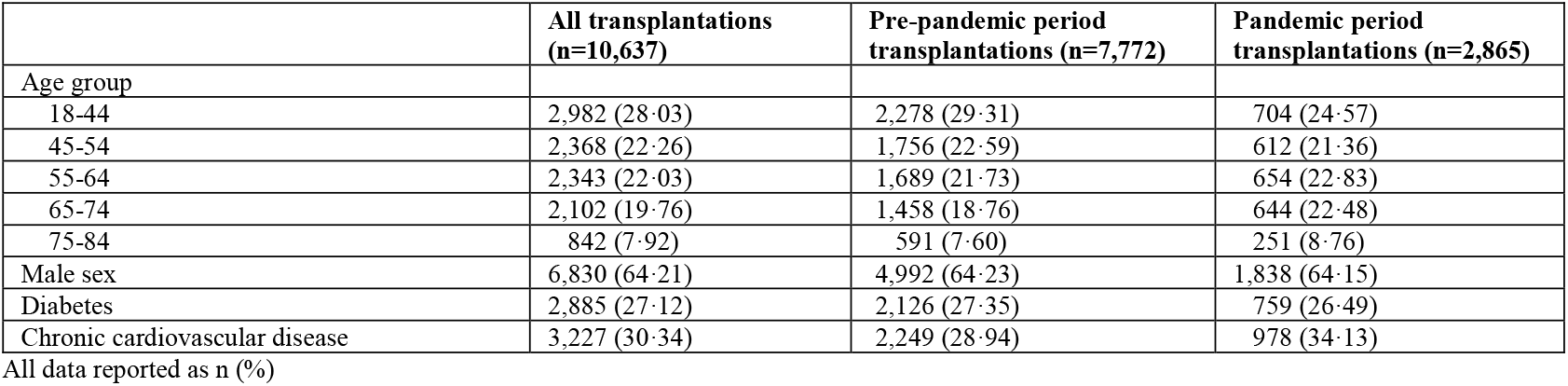
Characteristics of the kidney transplant recipients included in the study.

Transition flows along the follow-up of the 10,637 KTR of the study (Figure 3) indicate that during the 15,115 person-years followed up in the pre-pandemic period, 324 death events were observed, while during the 14,657 person-years followed-up in the pandemic period, 430 death events were observed, including 127 deaths observed among the 659 KTR hospitalized for COVID-19. The upper panel of Figure 4 shows how the cumulative probability of death during the pre-pandemic and pandemic periods evolved according to time since transplantation date. The crude HR [95% confidence interval] of death during pandemic period versus pre-pandemic period was 1·59 [1·37–1·86] globally, 1 ·78 [1·42–2·23] when restricted to the first year after transplantation, and 1 ·45 [1·18– 1·78] when considering only the following years (Table 2, model M1). However, the excess mortality was concentrated in KTR with COVID-19-related hospitalization (red curve in the bottom panel of Figure 4). Conversely, during the pandemic period, survival of KTR without COVID-19-related hospitalization (mauve curve in the bottom panel of Figure 4) was similar to that of the KTR during the pre-pandemic period (green curve in the bottom panel of Figure 4).

**Table 2.** Associations of pandemic period with mortality in kidney transplant recipients, univariable model and model adjusted for history of COVID-19-related hospitalization.

| Model | Variable | Variable value | Time range |  |  |  |  |  |  |  |  |
| --- | --- | --- | --- | --- | --- | --- | --- | --- | --- | --- | --- |
|  |  |  | From transplantation to study end |  |  | First year after transplantation |  |  | From the beginning of the second year after transplantation until study end |  |  |
|  |  |  | Deaths observed (n) / person-years(n) | HR [95% CI] | P | Deaths observed (n) / person-years (n) | HR [95%CI] | P | Deaths observed (n) / person-years (n) | HR [95% CI] | P |
| M1: unadjusted model | Period | Pre-pandemic (reference) | 324/15,115 | 1 (reference) |  | 172/6,598 | 1 (reference) |  | 152/8,517 | 1 (reference) |  |
|  |  | Pandemic | 430/14,657 | 1.59 [1.37–1.86] | <0.01 | 139/3,110 | 1.78 [1.42–2.23] | <0.01 | 391/11,547 | 1.45 [1.18–1.78] | <0.01 |
| M2: adjusted for history of COVID-19-related hospitalization | Period | Pre-pandemic (reference) | 324/15,115 | 1 (reference) |  | 172/6,598 | 1 (reference) |  | 152/8,517 | 1 (reference) |  |
|  |  | Pandemic | 430/14,657 | 1.15 [0.97–1.35] | 0.09 | 139/3,110 | 1.28 [1.00–1.65] | <0.05 | 391/11,547 | 1.05 [0.84–1.30] | 0.09 |
|  | COVID-19-related hospitalization | None (reference) | 303/14,208 | 1 (reference) |  | 97/3,010 | 1 (reference) |  | 206/11,198 | 1 (reference) |  |
|  |  | ≥1 hospitalization | 127/449 | 13.59 [11.15–16.57] | <0.01 | 42/100 | 13.66 [9.51–19.63] | <0.01 | 85/349 | 13.56 [10.71–17.18] | <0.01 |

**Figure 3.**
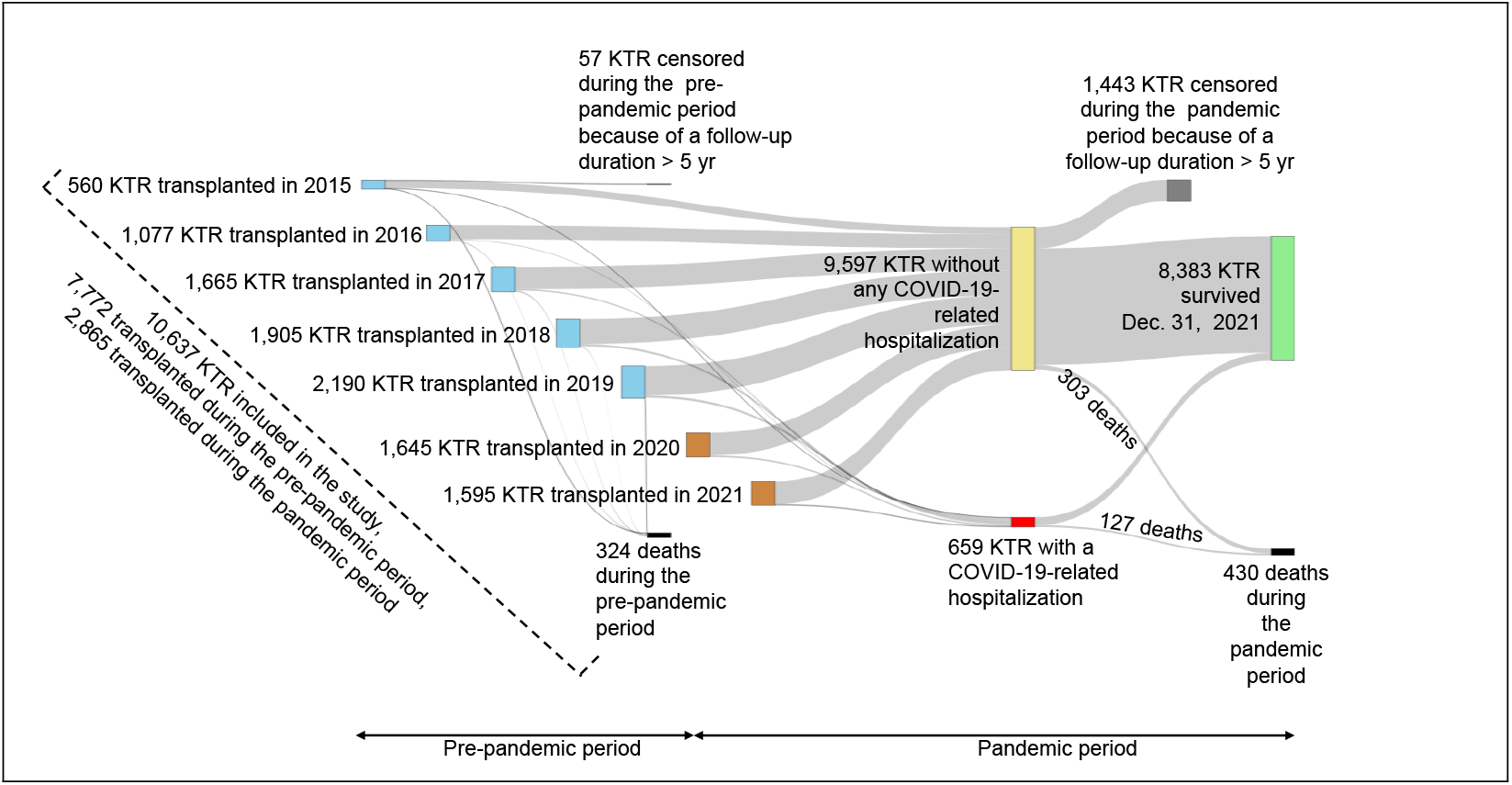
Detailed follow-up of KTR in the pre-pandemic and pandemic periods.

**Figure 4.**
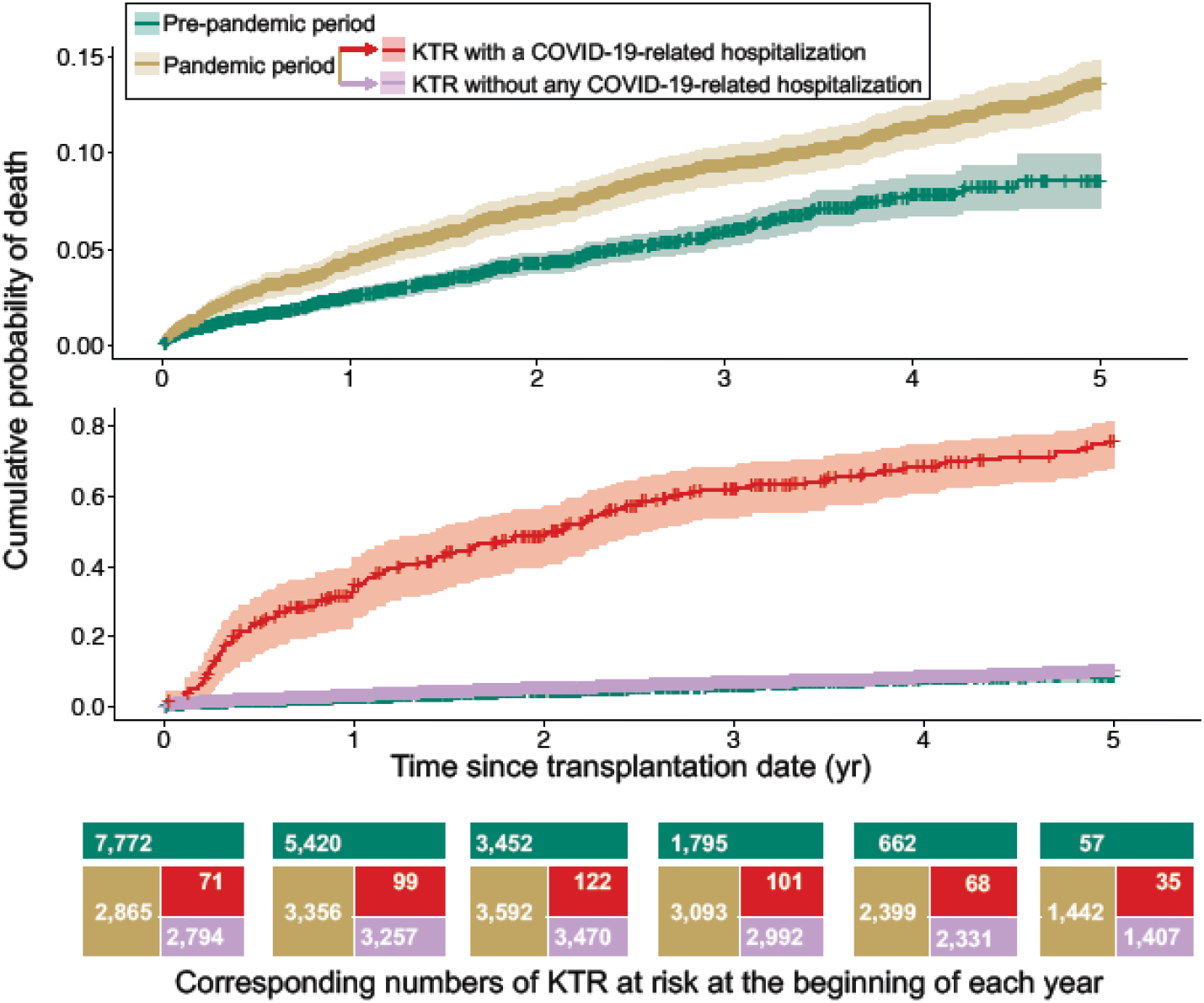
Probability of death according to time since transplantation date. Kidney transplant recipients followed-up during the pandemic period are shown altogether in the upper panel (pale gold curve), and split in the bottom panel according to whether they experienced a COVID-19-related hospitalization (red curve) or not (mauve curve). The pre-pandemic curve (green curve) is shown in both panels.

This finding is supported by the multivariable analysis when considering whether individuals experienced COVID-19-related hospitalization (Table 2, model M2): when adjusting for this factor, the risk of death during the pandemic period was not different from that during the pre-pandemic period (HR [95% CI] = 1 ·15 [0·97–1·35]), while in contrast, experiencing COVID-19-related hospitalization was associated with a dramatic higher risk of death (HR = 13·59 [11·15–16·57]). Therefore, the analysis of model M2 suggests that the excess mortality during the pandemic period in the univariable model M1 was primarily driven by the KTR with COVID-19-related hospitalizations. Nevertheless, in the adjusted model M2, an excess risk of death remains observed in KTR experiencing the pandemic during the first year after their transplantation date (HR = 1·28 [1·00–1·97]), while the very high risk of death in individuals with COVID-19-related hospitalization was stable whether considering the first year after the transplantation date or considering the following years (Table 2, model M2).

Table 3 presents the HR issued from a third model (model 3) which extends the adjustment made in model 2 to additional relevant covariates: as observed with model 2, the risk of death during the pandemic period was similar to that observed during the pre-pandemic period, and experiencing COVID-19-related hospitalization dramatically increased the risk of death. Model 3 further indicates that the risk of death was higher in the presence of chronic diseases, and this risk also increased with age. The risk of death was not associated with sex or dialysis duration, whereas pre-emptive transplantation was associated with a higher post-transplantation survival (HR 0·71 [0·56– 0·89]). Additional analyses presenting the dynamics of COVID-19 vaccination and investigating the impact of vaccination are shown in the Supplementary Appendix: landmark analyses detail how the course of vaccination status in KTR was associated with the probability of hospitalization for COVID-19. The Supplementary Appendix also shows that adjusting for the vaccination status rather than for a history of hospitalization in the Cox model did not modify the patterns of the HRs estimated in model 3, except for dialysis duration before transplantation that became associated with the risk of death. While the first and second doses of vaccine were not associated with the risk of death, receiving three doses or more was associated with a 58% decrease in the risk of death (HR 0·42 [0·30–0·58]).

**Table 3.** Multivariable analysis of factors associated with mortality in kidney transplant recipients (model 3)

| Variable | Variable value | Time range |  |  |  |  |  |  |  |  |
| --- | --- | --- | --- | --- | --- | --- | --- | --- | --- | --- |
|  |  | From transplantation to study end |  |  | First year after transplantation |  |  | From the beginning of the second year after transplantation until study end |  |  |
|  |  | Deaths observed (n) / person-years (n) | HR [95% CI] | P | Deaths observed (n) / person-years (n) | HR [95% CI] | P | Deaths observed (n) / person-years (n) | HR (95% CI) | P |
| Period | Pre-pandemic (reference) | 324/15,115 | 1 (reference) |  | 172/6,598 | 1 (reference) |  | 152/8,517 | 1 (reference) |  |
|  | Pandemic | 430/14,657 | 0.94 [0.78–1.12] | 0.46 | 139/3,110 | 1.09 [0.83–1.42] | 0.54 | 391/11,547 | 0.83 [0.66–1.05] | 0.12 |
| Hospitalization for COVID-19 | None (reference) | 303/14,208 | 1 (reference) |  | 97/3,010 | 1 (reference) |  | 152/8,517 | 1 (reference) |  |
|  | 1 hospitalization | 100/373 | 10.66 [9.61–13.38] | <0.01 | 29/80 | 9.75 [6.43–14.82] | <0.01 | 71/293 | 11.26 [8.58–14.78] | <0.01 |
|  | ≥2 hospitalizations | 27/76 | 14.31 [9.61–21.31] | <0.01 | 13/20 | 19.24 [10.63–34.82] | <0.01 | 14/56 | 11.35 [6.57–19.61] | <0.01 |
| Age group | 18–44 | 48/9,129 | 1 (reference) |  | 17/2,809 | 1 (reference) |  | 31/6,320 | 1 (reference) |  |
|  | 45–54 | 79/6,972 | 1.94 [1.35–2.77] | <0.01 | 37/2,201 | 2.53 [1.42–4.49] | <0.01 | 42/4,771 | 1.59 [1.00–2.53] | <0.05 |
|  | 55–64 | 163/6,458 | 3.62 [2.61–5.01] | <0.01 | 60/2,137 | 3.58 [2.07–6.17] | <0.01 | 103/4,321 | 3.60 [2.39–5.41] | <0.01 |
|  | 65–74 | 277/5,265 | 6.87 [5.02–9.42] | <0.01 | 121/1,837 | 7.84 [4.67–13.19] | <0.01 | 156/3,428 | 6.25 [4.20–9.30] | <0.01 |
|  | 75–84 | 187/1,948 | 11.90 [8.60–16.47] | <0.01 | 76/724 | 11.30 [6.60–19.34] | <0.01 | 111/1,224 | 12.46 [8.28–18.76] | <0.01 |
| Sex | Female | 243/10,683 | 1.09 [0.93–1.27] | 0.30 | 103/3,476 | 1.10 [0.87–1.40] | 0.43 | 140/7,276 | 1.06 [0.86–1.29] | 0.59 |
| Diabetes | Yes | 409/9,656 | 1.36 [1.17–1.58] | <0.01 | 151/2,880 | 1.29 [1.02–1.62] | 0.03 | 258/6,776 | 1.43 [1.18–1.75] | <0.01 |
| Chronic cardiovascular disease | Yes | 471/9,672 | 1.90 [1.62–2.23] | <0.01 | 183/3,047 | 1.85 [1.45–2.35] | <0.01 | 288/6,625 | 1.94 [1.57–2.39] | <0.01 |
| Dialysis duration (year) |  | 754/29,772 | 1.04 [0.97–1.11] | 0.30 | 211/9,708 | 1.02 [0.93–1.12] | 0.67 | 543/20,064 | 1.05 [0.86–1.17] | 0.41 |
| Pre-emptive kidney transplant | Yes | 120/8,510 | 0.71 [0.56–0.89] | <0.01 | 39/2,324 | 0.73 [0.49–1.07] | 0.10 | 81/6,186 | 0.69 [0.51–0.92] | 0.01 |

## Discussion

The study reported here addresses four main issues considering the whole French subpopulation of persons who benefited from a kidney transplant since year 2015: the indirect impact of the pandemic period on the survival of persons with no severe COVID-19-related event, the impact of experiencing COVID-19-related hospitalizations, the course and impact of vaccination, and the global relationship between various relevant factors and survival.

First, to our knowledge, this study is the first to investigate the indirect impact of the pandemic on the health status of KTR. Indeed, several features specific to the pandemic period such as the prioritization of hospital beds for patients with severe COVID-19^12,21^ and modifications of immunosuppressive regimen ^22^ have contributed to modify the standard care management of KTR. Such modifications might result in decreasing the long-term kidney function and overall survival of KTR, a feature not yet detectable during the limited follow-up in our study. However, the present study demonstrates that no excess mortality was yet observed during the pandemic period (bottom panel of Figure 4, model 3 in Table 3) in the whole national French sub-population of persons having benefited from a kidney transplant since 2015 and with no history of COVID-19-related hospitalization. Such an important result indicates that despite the disruptions of the French healthcare system related to the pandemic, the survival of the persons living with a kidney transplant has not been worsened.

Nevertheless, this result contrasts with the dramatic increase in mortality observed when considering the remaining individuals studied, i.e., the KTR who experienced COVID-19-related hospitalizations. Unfavourable outcomes in KTR hospitalized for COVID-19 when compared to persons without solid organ transplant have been previously reported.^23,24^ However, these results differ with those reported in other studies based on propensity-matched cohort analyses.^25–27^ Our large-scale study further quantifies the excess mortality specifically related to such hospitalizations in KTR when compared to KTR not hospitalized for COVID-19 at the national level (bottom panel of Figure 4, model 2 in Table 2, and model 3 in Table 3). Our analyses indicate that the development of a severe COVID-19 in KTR dramatically worsened their prognosis, as compared to those who did not experience such COVID-19-related hospitalizations.

Third, the present study details the dynamics of the COVID-19 vaccination in KTR at the national level, and investigates how vaccination has impacted COVID-19-related hospitalizations and survival (see Supplementary Appendix). In our study, a second and a third vaccine dose were associated with 35% and 67% decreases in the risk of COVID-19-related hospitalization, respectively. A Danish study reported that a second dose of vaccine was not associated with a lower risk of hospitalization. ^28^ A recent French study showed a strong association between immunosuppressant consumption and being hospitalized with COVID-19 after a full vaccination. ^11^ We found that the first and second doses of vaccine were not associated with the risk of death, while a third dose or more was associated with a 58% decrease in the risk of death. Since most of the KTR who were vaccinated received two first doses of BNT162b2 (see Supplementary Appendix), our results are in agreement with those of a national study on solid organ transplant recipients in England which reported a similar absence of association between two vaccine doses of BNT162b2 and the risk of death (HR 0 ·97 [0·71–1·31]).^29^ Moreover, our study further indicates that additional dose(s) of BTN162b2 were associated with lower risks of COVID-19-related hospitalization and death in the vulnerable population of KTR, and advocates for an enhanced vaccination process (schedule and doses) in KTR whose immunosuppressant consumption increases risks of hospitalization and death.

Fourth, the investigations globally reported here do not only concern the pandemic but estimate the whole relationship of relevant covariates with the survival of KTR. The information on the association of other covariates (age, sex, diabetes, cardiovascular comorbidities) with mortality should be considered as additional valuable side results. Importantly, pre-emptive transplantation was associated with a 30% lower risk of death and dialysis duration before transplantation in the remaining KTR was not associated with the risk of death (see model 3 in Table 3), in contrast with other studies reporting that dialysis duration before the transplantation was associated with a higher risk of death. ^30,31^ Two features have likely contributed to our study results. First, adopting a design based on the G9 mapping inherently resulted in selecting a target sub-population of relatively recent incident KTR (median follow-up: 3.4 years) with a time spent on dialysis < 5 years. Second, there is an overwhelming effect of hospitalizations for COVID-19 on mortality. Such an hypothesis is supported by the analysis shown in the Supplementary Appendix: when hospitalization for COVID-19 was removed from the model, dialysis duration was associated with a higher risk of mortality (9% per year).

Our study has several limitations. The first one is inherent to the SNDS data: the study design implied that only incident ESKD patients between January 1, 2015 and December 31, 2020 nationwide and who received a transplant between January 1, 2015 and December 31, 2021 were included, and therefore KTR who had spent more than 5 years on dialysis were not considered in this study. Moreover, the design inherently implied longer durations of dialysis in KTR transplanted at recent dates, with potential resultant confounding when studying the association of the pandemic period with mortality. However, the study design allowed us to consider factors such as the mode of entry into the transplant, the duration of dialysis and retrieve comorbidities with G9 mapping, including the incidence of comorbidities which are updated every year. Another limitation of the study is due to its observational nature which raises usual critical concerns. We tried to mitigate as much as possible the inherent flaws of this observational study by taking into account time-dependence of covariates, adjusting the estimates with features that we deemed relevant, and we applied STROBE recommendations for reporting. ^14^ Nevertheless, as underlined by others,^3,4^ exposure to the different variants of the virus, mitigation measures, health care system reorganization, vaccination schedules, and unknown confounding factors likely varied from one setting to another and with time, while additional methodological issues relating to the data collected and reported (e.g., various biases, relatively limited sample sizes) were not rare, globally yielding most caution about the generalizability of studies’ findings. In regards to generalizability issues, the main lines of the methodological framework proposed in the present study could be replicated by others for investigating the impact of the pandemic on other diseases and in other countries. One may consider that using COVID-19-related hospitalization as a proxy of severe COVID-19 episodes is another limit of the study. Direct COVID-19-related deaths that potentially occurred at home with no corresponding COVID-19-related hospitalization might have resulted in underestimating the direct COVID-19-related deaths in the analyses. Nevertheless, whatever the death cause and death place, all death events that occurred during the pre-pandemic and pandemic periods were included and appropriately handled in the analyses comparing the survival of KTR during the pre-pandemic and pandemic periods.

This study has several strengths. It was conducted at the national level of a European country with more than 66 million inhabitants, and more than 10,000 KTR totalizing nearly 30,000 person-years were followed-up. In addition, basing analyses on recent data allowed considering the main waves of the pandemic and including investigations on the impact of a third vaccine dose.

In conclusion, this national observational study showed a high excess mortality during the pandemic period in KTR with COVID-19-related hospitalization. However, in contrast with this dramatic direct impact of the disease, no indirect impact of the pandemic period on the survival of KTR was detected during study follow-up. The study results further indicate that a third dose or more of vaccine was associated with a reduced risk of death in the vulnerable population of KTR.

## Data Availability

According to the principles of data protection and French regulations, the authors cannot publicly release the data from the French National Health Data System (SNDS). However, any person or structure, public or private, for- profit or non-profit, can access SNDS data upon authorisation from the French Data Protection Office

## Acknowledgements

The COVID-HOSP working group: Tristan Delory, Centre Hospitalier Annecy Genevois, Annecy, France; Fanny Duchaine, IRDES, Paris, France; Maude Espagnacq, IRDES, Paris, France; Coralie Gandré, IRDES, Paris, France; Gilles Hejblum, INSERM, Paris, France; Myriam Khlat, INED, Aubervilliers, France; Nathanaël Lapidus, INSERM, Paris, France; Sophie Le Cœur, INED, Aubervilliers, France; Elhadji Leye, INSERM, Paris, France; Paul Moulaire, INSERM, Paris FRANCE; Jonas Poucineau, INED, Aubervilliers, France.

This work was supported by the Initiative Économie de la Santé of Sorbonne Université (Idex Sorbonne Universite’, programmes Investissements d’Avenir), and by the Ministère de la Solidarité et de la Santé (PREPS 20-0163). The sponsor and the funders had no role in study design, data collection and analysis, decision to publish, or preparation of the manuscript.

## Ethics statement

The National Health Data System (SNDS) is a set of strictly anonymous databases comprising all mandatory national health insurance reimbursement data. Since June 30, 2021 INSERM has direct access to the SNDS. This permanent access is given according French Decree No. 2016-1871 of December 26, 2016 relating to the processing of personal data called the “National Health Data System” and French law articles Art. R. 1461-1325 and 14. This study was declared prior to its initiation at the CépiDc-INSERM registry of studies requiring the use of the SNDS. In accordance with national legislation, written informed consent for participation was not required for this study.

## Data sharing statement

According to the principles of data protection and French regulations, the authors cannot publicly release the data from the French National Health Data System (SNDS). However, any person or structure, public or private, for- profit or non-profit, can access SNDS data upon authorisation from the French Data Protection Office (CNIL Commission Nationale de l’Informatique et des Libertés) to carry out a study, research, or an evaluation of public interest (https://www.snds.gouv.fr/SNDS/Contexte-et-perspectives-reglementaires#).

## Author Contributions

TD, GH, and NL initiated the study. GH and NL supervised the study; KEK, GH, NL, and EL designed the experimental plan; EL managed data and performed the analyses; GH, NL, and EL can take responsibility for the integrity of the data and the accuracy of the data analysis, EL is the guarantor; GH, NL, and EL prepared the first draft of the manuscript; All authors (TD, ME, KEK, GH, MK, NL, SL and EL) contributed to interpretation of the data, critically revised the manuscript, and approved the final version.

