## supplementary for "Direct and indirect impact of the COVID-19 pandemic on the survival of kidney transplant recipients: a national observational study in France" (from another version of this preprint: version 1, 2023-04-06)

### Dynamics and impact of COVID-19 Vaccination in kidney transplant recipients (KTR).

Figure S1 illustrates the dynamics of vaccination in the 10,141 KTR who were alive at the roll-out of the COVID-19 vaccination. On March 31, 2021, 3 months after the beginning of the COVID-19 vaccination roll-out in France (December 27, 2020) 60% of KTR had received at least one dose of vaccine while after one year, on December 31, 2021, 87% of KTR had received at least 2 doses and 71% had received a third dose or more. Among the 8,898 KTR who received two doses or more, 7,537 (85%) had received two first doses of Bnt162b2 (Pfizer-BioNTech mRNA vaccine), and among the 7,208 KTR who received three doses or more, 5,932 (82%) had received three first doses of Bnt162b2.

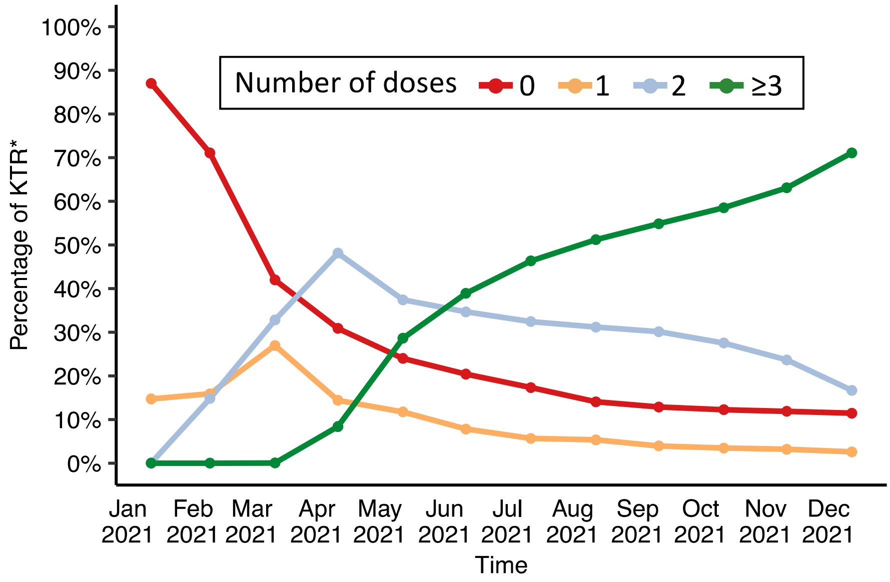


*The denominator used to calculate the percentages is the number of KTR who were included in the study and were alive on December 27, 2020, the date of the beginning of the vaccination roll-out in France: N = 10,141. Abbreviation used: KTR, kidney transplant recipient.

**Figure S1. Dynamics of COVID-19 vaccination in kidney transplant recipients (KTR) included in the study**

We investigated the risk of COVID-19-related hospitalization in KTR according to the vaccination schedule. The corresponding analyses (Figure S2 and Table S1) were restricted to the KTR who were at risk at the beginning of the pandemic period, i.e., the 7,448 KTR included in the study who had a transplantation before March 1, 2020. Panel A of Figure S2 shows how the probability of COVID-19-related hospitalizations evolved with time ($t_0$ is the pandemic period beginning, March 1, 2020, $t_{22}$ is the end-point date of the study, December 31, 2021) according to the cumulative number of vaccine doses received. The corresponding adjusted hazard ratio (HR) [95% confidence interval] of COVID-19-related hospitalization was respectively 0·62 [0·40–0·96], 0·63 [0·46–0·87] and 0·31 [0·21–0·46] for a first, second and third dose versus none (Table S1). The corresponding model S1 further indicates that the risk of COVID-19-related hospitalization was increased in the presence of chronic diseases. Sex and age were not associated with COVID-19-related hospitalization while dialysis duration was associated with an increased risk of COVID-19-related hospitalization. The vaccine roll-out started 10 months after the beginning of pandemic (Figure S2, panel A), a lag period during which there were no counterfactual observations. In order to get more insight on the effect of vaccination schedule on hospitalizations, we conducted 3 additional landmark analyses:

- The first landmark analysis began on the date when the first person in KTR of the study received his/her first dose, January 5, 2021 (Figure S2, panel B);
- The second landmark analysis began on the date when the first person in KTR of the study received his/her second dose, January 25, 2021 (Figure S2, panel C);
- The third landmark analysis began on the date when the first KTR of the study received his/her third dose, March 5, 2021 (Figure S2, panel D).

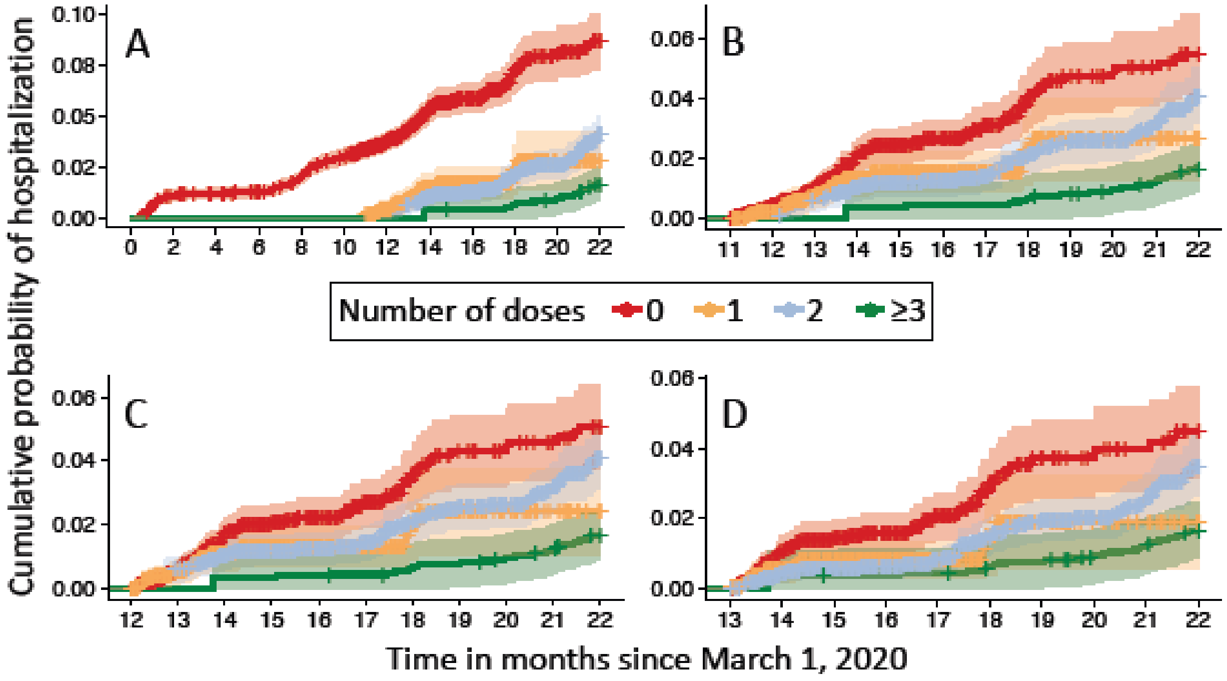


Panel A: cumulative probability of hospitalization according to the number of vaccine doses. Panel B: the first landmark analysis began on the date when the first KTR of the study received his/her first dose, January 5, 2021. Panel C: the second landmark analysis began on the date when the first KTR of the study received his/her second dose, January 25, 2021. Panel D: the third landmark analysis began on the date when the first KTR of the study received his/her third dose, March 5, 2021.

**Figure S2. Landmark analyses of the cumulative probability of COVID-19-related hospitalization according to the time-dependent cumulative number of doses**

As shown in Table S1, the patterns of the HRs for vaccination observed in the global analysis were not modified in the landmark analyses.

We investigated the association between vaccination schedule and deaths. Adjusted HR from the multivariable model S2 are reported in Table S2. Compared with model 3 shown in the main manuscript (see Table 3), model S2 was adjusted for vaccination and not for hospitalization: hospitalization and vaccination cannot be simultaneously considered in a model examining mortality, as the occurrence of severe COVID-19, of which COVID-19-related hospitalization is a proxy, mediates the association between vaccination and death. The pandemic period was associated with a higher risk of death (HR 1·36 [1·14–1·64]). This increase especially concerned the follow-up of KTR during their first year after transplantation (HR 1·61 [1·24–2·10] versus 1·21 [0·95–1·20] for the following years). Furthermore, while the first and second doses of vaccine were not associated with mortality, receiving 3 doses or more was associated with a much lower risk of death (HR 0·42 [0·30–0·58]). Model S2 also indicates that the risk of death was higher in the presence of chronic diseases, and this risk also increased with age, while the risk of death was similar in men and women. Dialysis duration was associated with a higher risk of death (HR 1·09 [1·02–1·17]), whereas pre-emptive transplantation was associated with a lower risk of death (HR 0·73 [0·58–0·92]).

| Variable | Variable value | Time range | | | | | | | |
|---|---|---|---|---|---|---|---|---|---|
| | | **From the beginning of pandemic period until study end** | | **From the date on which the first person of the study received the first dose (January 5, 2021) until study end** | | **From the date on which the first person of the study received the second dose (January 25, 2021) until study end** | | **From the date on which the first person of the study received the third dose (March 5, 2021) until study end** | |
| | | **Hospitalizations observed (n) / person-years (n)** | **HR [95% CI]** | **Hospitalizations observed (n) / person-years (n)** | **HR [95%CI]** | **Hospitalizations observed (n) / person-years (n)** | **HR [95% CI]** | **Hospitalizations observed (n) / person-years (n)** | **HR [95% CI]** |
| Vaccine against COVID-19 | 0 | 348/8,232 | 1 (reference) | 102/1572 | 1 (reference) | 78/1128 | 1 (reference) | 56/814 | 1 (reference) |
| | 1 | 26/590 | 0·62 [0·40–0·96] | 26/590 | 0·62 [0·40–0·96] | 22/473 | 0·63 [0·39–1·02] | 14/339 | 0·56 [0·31–1·01] |
| | 2 | 79/1,785 | 0·63 [0·46–0·87] | 79/1,785 | 0·65 [0·47–0·89] | 79/1,785 | 0·67 [0·48–0·93] | 69/1,627 | 0·60 [0·42–0·86] |
| | ≥3 | 56/2,338 | 0·31 [0·21–0·46] | 56/2,338 | 0·32 [0·22–0·48] | 56/2,338 | 0·33 [0·22–0·50] | 56/2,338 | 0·31 [0·20–0·46] |
| Age group | 18–44 | 110/3,784 | 1 (reference) | 58/1,754 | 1 (reference) | 56/1,585 | 1 (reference) | 50/1,516 | 1 (reference) |
| | 45–54 | 113/3,042 | 1·24 [0·95–1·62] | 61/1,455 | 1·29 [0·90–1·86] | 51/1,316 | 1·13 [0·77–1·65] | 41/1,210 | 1·09 [0·71–1·65] |
| | 55–64 | 139/2,888 | 1·45 [1·12–1·88] | 70/1,414 | 1·41 [0·99–2·02] | 60/1,279 | 1·27 [0·87–1·85] | 48/1,136 | 1·26 [0·84–1·90] |
| | 65–74 | 101/2,352 | 1·21 [0·91–1·61] | 53/1,180 | 1·21 [0·82–1·79] | 50/1,075 | 1·19 [0·79–1·78] | 43/921 | 1·36 [0·89–2·09] |
| | 75–84 | 46/879 | 1·51 [1·05–2·16] | 20/482 | 1·14 [0·67–1·95] | 18/442 | 1·05 [0·60–1·85] | 13/335 | 1·06 [0·55–2·03] |
| Sex | female | 166/4,689 | 0·89 [0·73–1·07] | 97/2,287 | 1·05 [0·81–1·35] | 89/2,074 | 1·10 [0·84–1·43] | 77/1,861 | 1·18 [0·88–1·57] |
| Diabetes | Yes | 254/4,452 | 1·63 [1·35–1·96] | 131/2,128 | 1·79 [1·38–2·31] | 115/1,927 | 1·71 [1·30–2·25] | 97/1,727 | 1·80 [1·33–2·42] |
| Chronic cardiovascular disease | Yes | 239/4,531 | 1·25 [1·03–1·51] | 114/2,169 | 1·14 [0·88–1·49] | 103/1,963 | 1·18 [0·89–1·56] | 85/1,760 | 1·21 [0·89–1·64] |
| Dialysis duration (year) | | 509/12,945 | 1·16 [1·07–1·24] | 263/6,285 | 1·16 [1·04–1·27] | 235/5,697 | 1·15 [1·03–1·28] | 195/5,118 | 1·05 [0·93–1·18] |

Abbreviations used: HR [95% CI], hazard ratio [95% confidence interval]

**Table S1. Multivariable analysis of factors associated with the risk of COVID-19-related hospitalization in kidney transplant recipients (model S1)**

| Variable | Variable value | Time range | | | | | | | | |
|---|---|---|---|---|---|---|---|---|---|---|
| | | From transplantation date to study end | | | First year after transplantation date | | | From the beginning of the second year after transplantation date until study end | | |
| | | Deaths observed (n) / person-years (n) | HR [95% CI] | P | Deaths observed (n) / person-years (n) | HR [95%CI] | P | Deaths observed (n) / person-years (n) | HR [95% CI] | P |
| Period | Pre-pandemic (reference) | 324/15,115 | 1 (reference) | | 172/6,598 | 1 (reference) | | 152/8,517 | 1 (reference) | |
| | Pandemic | 430/14,657 | 1·36 [1·14–1·63] | <0·01 | 139/3,110 | 1·61 [1·24–2·10] | <0·01 | 391/11,547 | 1·21 [0·95–1·20] | 0·12 |
| Vaccine against COVID-19* | 0 | 284/8,997 | 1 (reference) | | 97/2,025 | 1(reference) | | 187/6,972 | 1(reference) | |
| | 1 | 25/761 | 0·96 [0·63–1·45] | 0·86 | 7/170 | 0·80 (0·37–1·73] | 0·58 | 18/591 | 1·04 [0·64–1·69] | 0·87 |
| | 2 | 78/2,171 | 0·99 [0·77–1·28] | 0·96 | 23/447 | 0·90 [0·56–1·42] | 0·65 | 55/1,724 | 1·03 [0·76–1·37] | 0·84 |
| | ≥3 | 43/2,728 | 0·42 [0·30–0·58] | <0·01 | 12/468 | 0·42 [0·23–0·77] | <0·01 | 31/2,260 | 0·42 [0·29–0·62] | <0·01 |
| Age group | 18–44 | 48/9,129 | 1 (reference) | | 17/2,809 | 1 (reference) | | 31/6,320 | 1 (reference) | |
| | 45–54 | 79/6,972 | 1·98 [1·38–2·84] | <0·01 | 37/2,201 | 2·55 [1·44–4·54] | <0·01 | 42/4,771 | 1·65 [1·04–2·63] | 0·03 |
| | 55–64 | 163/6,458 | 3·74 [2·70–5·18] | <0·01 | 60/2,137 | 3·69 [2·13–6·35] | <0·01 | 103/4,321 | 3·75 [2·50–5·64] | <0·01 |
| | 65–74 | 277/5,265 | 6·94 [5·07–9·51] | <0·01 | 121/1,837 | 7·67 [4·56–12·90] | <0·01 | 156/3,428 | 6·48 [4·36–9·62] | <0·01 |
| | 75–84 | 187/1,948 | 12·23 [8·83–16·94] | <0·01 | 76/724 | 11·94 [6·98–20·43] | <0·01 | 111/1,224 | 12·52 [8·31–18·86] | <0·01 |
| Sex | female | 243/10,683 | 1·06 [0·91–1·24] | 0·43 | 103/3,476 | 1·10 [0·87–1·40] | 0·44 | 140/7,276 | 1·04 [0·85–1·27] | 0·70 |
| Diabetes | Yes | 409/9,656 | 1·41 [1·22–1·64] | <0·01 | 151/2,880 | 1·31 [1·03–1·65] | 0·02 | 258/6,776 | 1·49 [1·23–1·82] | <0·01 |
| Chronic cardiovascular disease | Yes | 471/9,672 | 1·93 [1·65–2·26] | <0·01 | 183/3,047 | 1·84 [1·45–2·35] | <0·01 | 288/6,625 | 2·00 [1·62–2·46] | <0·01 |
| Dialysis duration (year) | | 754/29,772 | 1·09 [1·02–1·17] | 0·01 | 211/9,708 | 1·07 [0·98–1·18] | 0·13 | 543/20,064 | 1·12 [1·00–1·25] | 0·04 |
| Pre-emptive kidney transplant | Yes | 120/8,510 | 0·73 [0·58– 0·92] | <0·01 | 39/2,324 | 0·75 [0·51– 1·11] | 0·15 | 81/6,186 | 0·73 [0·54– 0·98] | 0·03 |

*the vaccination covariate was handled as an ordinal variable. Abbreviations used: HR [95% CI], hazard ratio [95% confidence interval]

**Table S2· Multivariable analysis of factors associated with mortality in kidney transplant recipients (model S2)**
